# Experimental hypoxia to probe neuro-metabolic and vascular dysregulation in ME/CFS: a multimodal proof-of-concept MRI study

**DOI:** 10.64898/2026.08.10.26359935

**Authors:** Viola Bader, Katharina Estermann, Eva Niess, Tobias Zrzavy, Florian Fischmeister, Teresa Haider, Birgit Ludwig, Frederik Barkhof, Henk JMM Mutsaerts, Gregor Kasprian, Fabian Niess, Wolfgang Bogner, Kathrin Kollndorfer, Lukas Haider

## Abstract

**Background:** Myalgic Encephalomyelitis/Chronic Fatigue Syndrome (ME/CFS) is a poorly understood, debilitating multisystem condition. Converging evidence implicates impaired cellular bioenergetics, neuroinflammation and defective neurovascular coupling that may manifest as “virtual hypoxia” only under physiological stress.

**Methods:** We performed a single-session multimodal 3T MRI study combining brain volumetry, arterial spin labelling (ASL) and multivoxel proton magnetic resonance spectroscopy under normoxia and two controlled hypoxic challenges (oxygen saturation 87 ± 3%) in 26 ME/CFS patients and 27 age- and sex-matched healthy controls.

**Results:** After intracranial-volume normalization, patients showed a reduced brainstem volume (1.46 ± 0.14 vs. 1.55 ± 0.18 % of eTIV; p = 0.013, FDR-p = 0.039), whereas deep grey matter and whole-brain parenchymal fraction did not differ between groups. Whole-brain cerebral blood flow (CBF) rose under hypoxia in both groups (controls +4.8 ± 13.0%, patients +3.7 ± 11.7%), with greater initial inter-individual variability in patients (patient-to-control variance ratio up to 6.94; FDR-p = 0.001). Thalamic lactate-to-creatine (Lac/tCr) ratios increased with hypoxia in controls (FDR-p = 0.028) but were already elevated at normoxia in patients (0.171 vs. 0.135; FDR-p = 0.021) and did not rise further (FDR-p = 0.38). In exploratory analyses, patients showed exaggerated inverse coupling between thalamic total N-acetylaspartate (tNAA/tCr) and white-matter CBF.

**Conclusions:** These findings provide *in vivo* evidence of impaired neuro-metabolic and vascular adaptive capacity in ME/CFS, supporting the virtual hypoxia hypothesis and highlighting candidate imaging markers for stratification that warrant validation.

## Introduction

Myalgic Encephalomyelitis/Chronic Fatigue Syndrome (ME/CFS) is a multifactorial, debilitating disease defined by substantial reductions in function, post-exertional malaise (PEM), unrefreshing sleep, cognitive impairment, and/or orthostatic intolerance that collectively undermine quality of life and functional capacity^1^. Despite substantial disease burden and growing prevalence estimates, the underlying pathophysiology remains unresolved and lacks robust biomarkers to support diagnosis, stratification, and targeted therapy. In contrast, Multiple Sclerosis (MS), another complex, potentially progressive neuroinflammatory condition in which fatigue is a leading symptom, has been interrogated systematically over the past century. In MS, convergent evidence implicates inflammation-related mitochondrial injury, impaired bioenergetics, and downstream tissue energy failure as central mechanisms linked to symptoms and disability^2,3^. Whether similar energy constraints at the tissue level contribute to fatigue and other neurological features in ME/CFS is unknown.

The brain’s energetic demand renders it particularly vulnerable to mismatches between supply and demand: constituting only about 2% of body mass, the brain consumes roughly 20% of the body’s oxygen and lacks energy reserves^4^. The concept of virtual hypoxia, where increased metabolic demand or impaired substrate utilization produces a functional state of hypoxia despite nominal oxygen availability, has been proposed in MS and related disorders that share features with ME/CFS^5^.

In ME/CFS, convergent evidence points to impaired cellular bioenergetics^6,7^, neuroinflammation and immune activation^8–10^, and altered cerebrovascular control or hypo cerebral blood flow (CBF) under physiological stressors^11^. However, static measurements at rest may miss dynamic deficits in metabolic-vascular coupling that become apparent only under load. Physiologically controlled hypoxic challenges increase oxygen delivery demands and probe neurovascular reactivity (NVR) and metabolic flexibility in vivo. Advanced MRI can quantify complementary facets of brain physiology: ASL provides non-contrast CBF quantification and vascular reactivity^12^; proton magnetic resonance spectroscopy (^1^H-MRSI) provides markers of oxidative and glycolytic metabolism and neuronal integrity (e.g., glutamate, glutamine, lactate, tNAA)^13,14^; and structural MRI assessment of regional brain atrophy, including brainstem nuclei implicated in autonomic regulation and fatigue^15,16^.

Prior neuroimaging studies in ME/CFS have yielded heterogeneous findings across modalities, often without physiological perturbations^10,17–20^. We therefore implemented a single-session, multimodal MRI paradigm tailored to minimize participant burden and assessed the brain’s adaptive responses across three experimental conditions: normoxia followed by two consecutive normobaric hypoxia challenges. We hypothesized that, relative to matched controls, ME/CFS would show (i) brain atrophy; (ii) altered metabolism at rest and (iii) impaired reorganization of metabolic-perfusion coupling during hypoxia, consistent with reduced metabolic flexibility and virtual hypoxia^2,3^. Establishing reproducible, quantitative stress-response signatures could accelerate biomarker development and enable mechanism-based clinical trials.

## Methods

### Study Participants

Individuals were recruited from inpatient and outpatient services at the Medical University of Vienna. The cohort comprised 26 participants with a neurologist-confirmed diagnosis of ME/CFS, aged 18-65 years (**Figure 1**), and 27 age- and sex-matched healthy controls.

**Figure 1.**
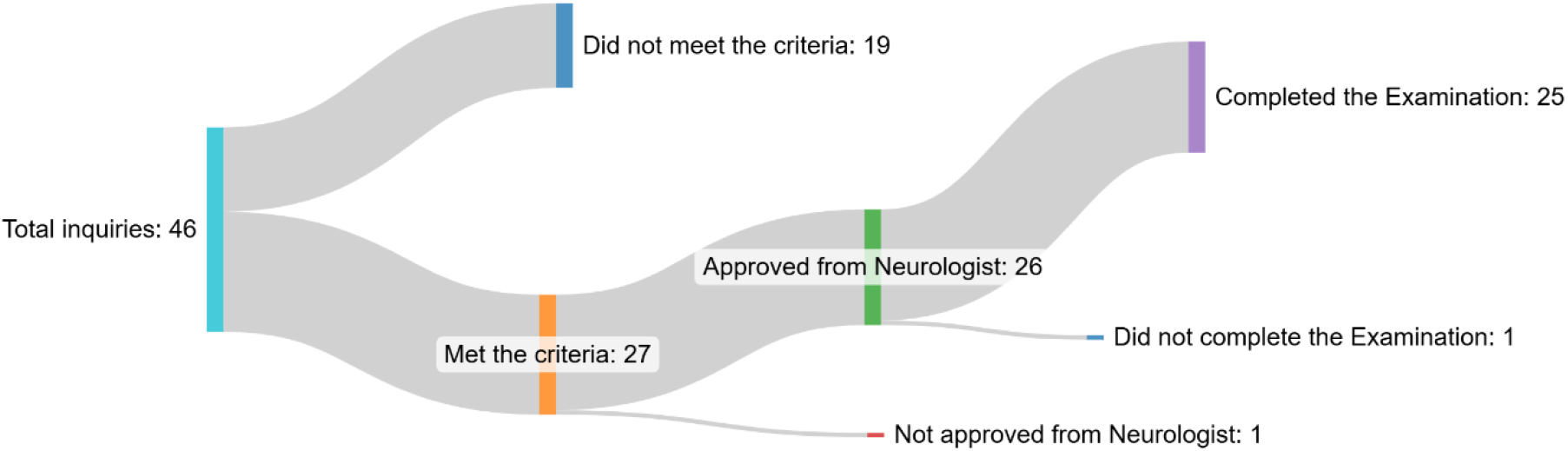
Recruitment flowchart for ME/CFS participants. From 46 total inquiries, 27 met the criteria, 26 were neurologist-approved, and 25 completed the examination.

Due to inconsistencies during scanning procedures, subsets of participants were excluded from specific analyses: All participants were included in brain volumetric analyses (ME/CFS: n=26; controls: n=27). For arterial spin labelling (ASL) CBF analyses, 4 ME/CFS participants and 11 controls were excluded, yielding final samples of ME/CFS: n=22 and controls: n=16. For magnetic resonance spectroscopic imaging (MRSI) analyses, 5 ME/CFS participants and 12 controls were excluded, yielding final samples of ME/CFS: n=21 and controls: n=15. Exclusions were due to incorrect acquisition order of normoxia and hypoxia sessions (controls/patients: 6/1), aborted examinations (3/2), corrupted datafiles (ASL: 2/1; MRSI: 1/1), and incorrect MRSI slice positioning (1/2).

### Ethical Considerations

The study was performed in accordance with the Declaration of Helsinki (1964), including current revisions. The study protocol was approved by the Ethics Committee of the Medical University of Vienna. All participants were informed about the aim of the study and gave written informed consent before inclusion.

### MR Protocol

All measurements were conducted at a clinical 3T MRI scanner (Magnetom Vida, Siemens Healthineers, Erlangen, Germany) using a 64-channel receiver head coil (Siemens Healthineers, Erlangen, Germany). All participants were measured in headfirst supine position.

High-resolution T_1_-weighted anatomical images were acquired using a MPRAGE sequence with spatial resolution: 0.98×0.98×1.00mm³, TR/TI/TE=2000/1010/3.26ms, 208 slices, GRAPPA factor 3, acquisition time (TA)=4:38min.

Cerebral CBF was measured using a background suppressed 3D gradient and spin echo (GRASE) pseudo-continuous arterial spin labelling (pCASL) sequence with the following parameters: spatial resolution: 3.4×3.4×4mm³ interpolated to 1.7×1.7×4mm³, TR=4000ms, TE=22.1ms, 32slices, post labelling delay=1800ms, labelling duration=1800ms. GRAPPA factor 2, 8 label/control pairs, TA=4:42min. Scans included a proton density M_0_ calibration image with identical geometry and TR, without labelling and without background suppression.

Metabolic imaging was performed using a 2D semi-LASER chemical shift imaging (CSI) spectroscopy sequence (matrix size=16×16, field of view (FOV)=160×160×15 mm³, volume of interest (VOI)=80×80×15mm³, TR=1700ms, TE=135ms, 3 averages, TA=∼8min). The CSI slice was positioned to cover the thalamus.

All subjects were first measured under normoxic conditions (MPRAGE, ASL, MRSI) followed by two consecutive sessions under hypoxia which included only ASL and MRSI scans, see **Figure 2a**. Hypoxic conditions (oxygen saturation 87±3%, ranging from 82-95%) were induced using a hypoxic generator (Longfian Scitech Co.).

**Figure 2.**
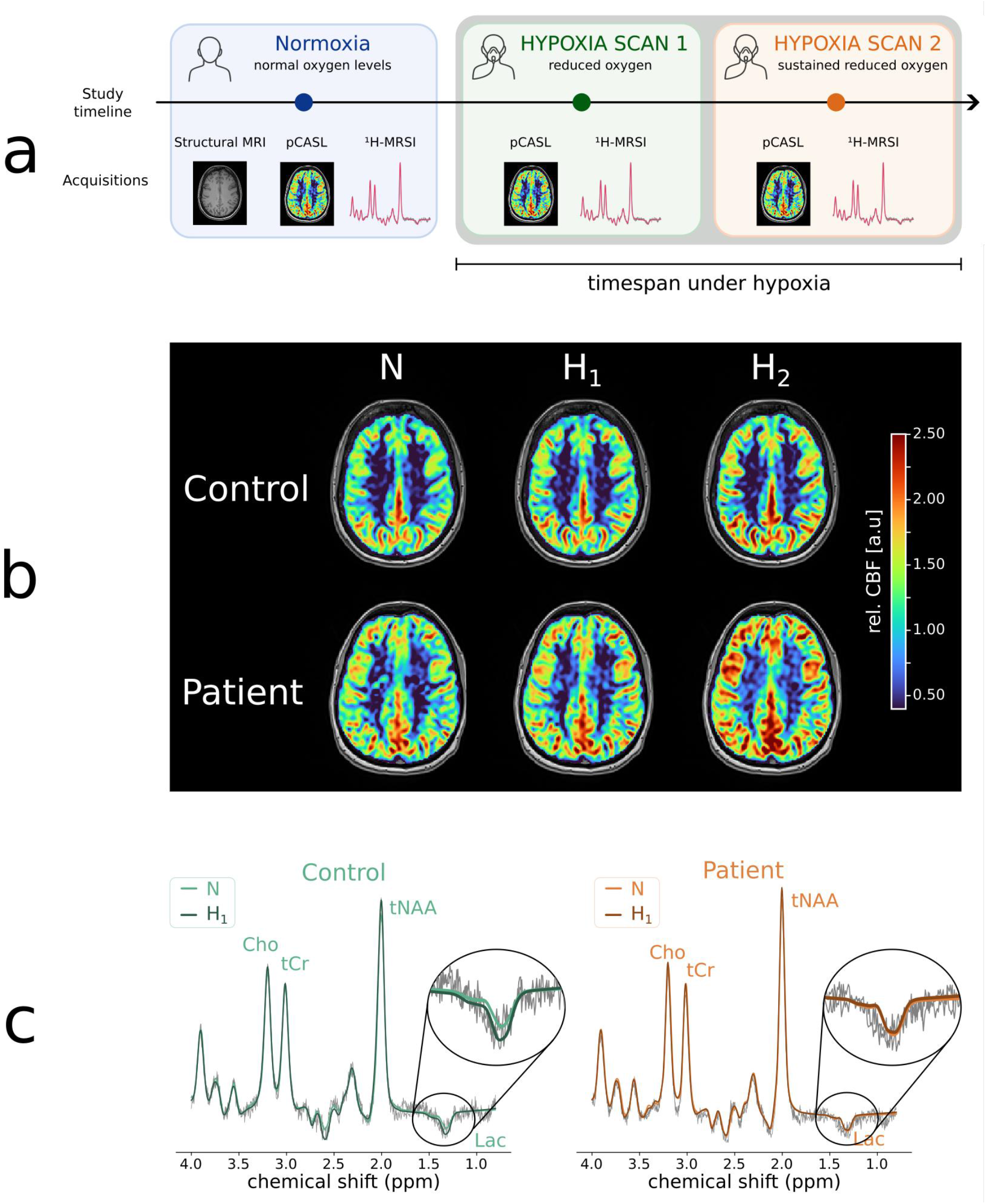
Experimental study design and exemplary cerebral blood flow maps and thalamic spectra. (a) Participants underwent MRI at baseline under normoxic conditions (normoxic scan=N) followed by two scans during hypoxic conditions (hypoxic scan 1 and 2=H1/H2), which were induced by controlled reduction of oxygen. At each condition, CBF imaging (TA=∼5min) and spectroscopic measurements (TA=∼8min) were acquired, structural MRI (TA=∼5min), used for regional volumetric analysis and anatomical localization were only acquired at the normoxia scan. Timespan spent under hypoxic condition was ∼30min and total scan time was below 1h. (b) Representative relative cerebral blood flow (CBF) maps acquired under normoxia (N) and hypoxic conditions (H1, H2) from a ME/CFS patient in their 30’s with a disease duration of more than 10 years and no preceding infection, and an age- and sex-matched healthy control. CBF maps are shown as relative CBF normalized to the mean whole-brain CBF of the corresponding normoxic baseline scan. Whole brain CBF increased during hypoxia in both subjects, with an increase of approximately 6% in the healthy control and 22% in the ME/CFS patient. (c) Representative overlaid thalamic spectra acquired under normoxic baseline conditions (N) and the first hypoxic challenge (H1) from a healthy control and a ME/CFS patient in their 50’s with a disease duration of less than 5 years. The inset shows a magnified view of the lactate (Lac) resonance. In healthy controls, Lac/tCr increased from N to H1, whereas the elevated baseline Lac/tCr observed in the ME/CFS patient remained largely unchanged during hypoxia. <u>Abbreviations</u>: MRSI: Magnetic Resonance Spectroscopy Imaging, CBF=cerebral blood flow; N=normoxia; H1=first hypoxic challenge; H2=second hypoxic challenge, Lac=lactate, tCr=total creatine, tNAA=total N-acetylaspartate, Cho=Choline, ppm=parts per million.

### MR Post-processing Analysis

Structural segmentation of T_1_-weighted MPRAGE images was performed using the automated FreeSurfer recon-all pipeline (version 7.4.1, https://surfer.nmr.mgh.harvard.edu). All segmentation outputs were transformed to structural space by registering to the corresponding T_1_-weighted images. WM, total GM consisting of cortical grey matter (GM), and deep GM (including thalamus, caudate nucleus, putamen, globus pallidus, hippocampus, amygdala, and nucleus accumbens), were obtained from FreeSurfer parcellations. For each ROI, volumes were estimated and later analyzed.

ASL images were processed and quantified using FSL’s BASIL toolbox^31^. Pre-processing included motion correction and pairwise subtraction of label and control images to generate CBF-weighted images, which were subsequently averaged. Quantitative CBF maps were estimated using a single-compartment kinetic model with voxel-wise calibration based on the M_0_ image. CBF maps were coregistered and upsampled to the native resolution of the T_1_-weighted images, and aligned with the FreeSurfer-derived masks. CBF maps were calculated as relative changes by normalizing each dataset to the corresponding mean whole brain normoxic CBF value. Regional CBF values were extracted for each region of interest.

Dicoms with coil-combined data were exported from the scanner and processed using an inhouse developed pipeline (MATLAB R2021, LCModel v6.3, Python v3.10). Prior to spectral fitting, voxel-wise B₀ correction was applied using LCModel-derived frequency shift maps to realign spectra, followed by zero-order phase correction to account for spatially varying phase offsets. Due to inherently low voxel-wise SNR, spectra were averaged within predefined regions of interest, and only voxels meeting quality criteria (FWHM ≤ 0.1ppm) were included in the analysis. After regional averaging, spectral fitting was performed using LCModel with a simulated basis set including 19 metabolites (alanine, ascorbic acid, creatine [Cr_3.0_/Cr_3.9_], GABA, glucose, glutamate, glutamine, glycine, glycerol-phosphocholine, glutathione, myoinositol, lactate, N-acetyl aspartate [NAA_singlet_/NAA_multiplet_], N-acetyl-aspartyl-Glutamate, phosphocholine, phosphocreatine [PCr_3.0_/PCr_3.9_], phosphorylethanolamine, scylloinositol, taurine). Metabolite levels in two regions of interest (ROIs) were analysed: white matter (WM)-dominated regions and the thalamus. WM masks were obtained using FSL FAST (threshold of 60%), while thalamus masks were obtained from FreeSurfer output. High-resolution masks were downsampled to the MRSI grid in k-space while accounting for point spread function and partial volume effects. Metabolite levels were quantified as ratios to total creatine (tCr).

### Questionnaires

The German version of the SF-36^32,33^ physical activity subscale was used to assess participants’ health limits. Activities ranging from washing and dressing to walking short and long distances, and even running, were performed during a typical day, and participants were asked to indicate whether they were limited a lot, a little, or not at all. The scores for the ten questionnaire items were transformed into a scale ranging from 0 to 100%, where higher scores indicate a better physical condition and lower scores indicate a worse condition.

Severity of chronic fatigue was assessed using the German version of the Chalder Fatigue Scale (CFQ)^34,35^. The 11-item CFQ is a brief screening instrument that can be used for epidemiological and clinical purposes. The abbreviation CFQ is commonly used to distinguish it from CFS. The scoring procedure of the instrument allows to differentiate between no fatigue and severe fatigue with an additional item to assess chronicity.

### Statistical Analysis

All statistical analyses were performed in Python (version 3.10) and R studio^36^.

Volumetric measures were normalized to estimated total intracranial volume (eTIV) to account for differences in brain size. Group effects on regional volumes were assessed using linear models with volume as dependent variable and group, age, sex, and eTIV as fixed effects (volume ∼ group + age + sex + eTIV). Group effects were corrected for multiple comparisons across the three tested regions (brain-stem, deep grey matter and whole-brain parenchymal fraction) using the Benjamini-Hochberg false-discovery-rate procedure.

Between-group differences in the inter-individual variability of the regional cerebral CBF response to hypoxia (relative CBF normalized to the normoxic baseline) were assessed separately for each hypoxic challenge (H_1_ and H_2_) by comparing patient-to-control variance ratios using the F-test, with 95% confidence intervals derived from 10,000 bootstrap resamples and p-values corrected across regions of interest using the Benjamini-Hochberg false-discovery-rate procedure.

Differences in cerebral blood flow (CBF) between conditions were evaluated separately for the whole brain, total GM, WM, and thalamus using linear mixed-effects models with CBF as the dependent variable and condition, group, age, and sex as fixed effects, including a random intercept for subjects (CBF ∼ condition + group + age + sex + (1|subject)). Additional models were computed, including clinical parameters as covariates (CBF ∼ condition + group + age + sex + clinical parameters + (1|subject)).

Spectroscopic data were analysed using metabolite ratios (e.g., Lac/tCr, tNAA/tCr, Cho/tCr) extracted from white matter and thalamus regions. Effects of condition and group were assessed using two linear mixed-effects models: the primary linear mixed-effects model included an interaction term (*Lac/tCr ∼ Condition × Group + age + sex [+ clinical parameters] + (1|Subject)*) and was evaluated with and without additional clinical parameters as covariates. Post hoc within-group analyses were performed by fitting separate linear mixed-effects models (*Lac/tCr ∼ Condition + age + sex + (1|Subject)*) for ME/CFS patients and controls. Baseline differences under normoxia (N) between groups were additionally evaluated using Welch’s *t*-test. Multiple-comparison correction was performed using the Benjamini–Hochberg false discovery rate (FDR) procedure, with p-values adjusted across the tested regions separately for each statistical analysis.

As an exploratory, hypothesis-generating analysis, multimodal associations were examined using correlation networks combining volumetric, spectroscopic, CBF, demographic and clinical variables. To mirror the reduced primary analyses and limit redundant comparisons, network nodes comprised eTIV-normalized brain-stem (including pons), deep grey-matter and whole-brain (parenchymal-fraction) volumes; thalamic and white-matter Lac/tCr and tNAA/tCr; total grey-matter, white-matter and thalamic perfusion; age; and SF-36 physical-functioning and Chalder fatigue scores. Whole-brain perfusion, which serves as the within-subject normalisation reference and is therefore invariant at normoxia, was retained in the primary perfusion analysis but not included as a network node. Within each group and condition, pairwise Spearman correlations were computed on complete cases; variables were grouped into modules by agglomerative average-linkage clustering of 1 - |r|, and the network was retained at |r| ≥ 0.5 following a threshold parameter sweep. Modules were defined by cutting the linkage dendrogram at a cophenetic distance of 0.5, equivalent to an average within-module |r| of 0.5. Reproducibility was quantified by subject-level bootstrap resampling (edge sign-consistency and module co-membership) instead of family-wise error correction, reflecting the exploratory aim, and differences from controls were summarised as a differential network (patient minus control correlations).

## Results

### Study design

All participants underwent in one session MRI under normoxia and hypoxia. Under normoxia we acquired T_1_-weighted anatomical images (MPRAGE), arterial spin labelling (pc-ASL), and magnetic resonance spectroscopy (^1^H-MRSI). To probe adaptive cerebrovascular and metabolic responses beyond resting conditions, the normoxic baseline examination (N) was followed by two controlled normobaric hypoxic challenges (H_1_ and H_2_), during which ASL and MRSI data were acquired. Normobaric hypoxia was induced immediately before the first hypoxic challenge by reducing the inspired oxygen fraction using a hypoxic generator, resulting in a mean peripheral oxygen saturation of 87±3% (range: 82-95%). Participants remained under hypoxic conditions throughout both hypoxic challenges (∼30min) while peripheral oxygen saturation was continuously monitored (**Figure 2a**). Representative cerebral blood flow (CBF) maps acquired under normoxia and both hypoxic challenges are shown in **Figure 2b**, illustrating the increase in CBF during hypoxia in both controls and patients. Representative overlaid thalamic spectra acquired under normoxia and the first hypoxic challenge are shown in **Figure 2c**, demonstrating the hypoxia-induced increase in Lac/tCr in a healthy control, whereas the elevated baseline Lac/tCr in the ME/CFS patient remained largely unchanged.

### Cohort characteristics

We enrolled 26 ME/CFS participants (18-65 years) and 27 age- and sex-matched healthy controls, detailed description presented in **Table 1**. Groups differed on SF-36 Physical Functioning and Chalder Fatigue total scores, as expected.

**Table 1.** Demographic and clinical characteristics of ME/CFS patients and healthy controls. <u>Abbreviations</u>: SF-36=Short Form Health Survey Physical Functioning score, CFQ=Chalder Fatigue Scale.

|  | Patients (n=26) | Control (n=27) | p-value |
| --- | --- | --- | --- |
| Age [years] | 42.1 ± 12.9 | 38.7 ± 11.4 | 0.31 |
| Sex [w/m] | 24 / 2 | 18 / 9 | 0.05 |
| Education | Lower secondary: 2<br>Apprenticeship/vocational school: 4<br>High school diploma: 11<br>University/university of applied sciences: 9 | Lower secondary: 1<br>Apprenticeship/vocational school: 6<br>High school diploma: 3<br>University/university of applied sciences: 16 | 0.20 |
| Income | <1000 €: 2<br>1000-<1500 €: 8<br>1500-<2000 €: 0<br>2000-<2500 €: 3<br>2500-<3000 €: 2<br>3000-<3500 €: 1<br>3500-<4000 €: 3<br>≥4000 €: 5<br>No answer: 2 | <1000 €: 0<br>1000-<1500 €: 0<br>1500-<2000 €: 0<br>2000-<2500 €: 1<br>2500-<3000 €: 1<br>3000-<3500 €: 2<br>3500-<4000 €: 1<br>≥4000 €: 18<br>No answer: 3 | <0.001 |
| Post-Infection Onset [yes/no] | 23 / 3 | - | - |
| Disease duration [years] | 6.3 ± 6.3 | - | - |
| Disease progression [rapid/insidious] | 7 / 19 | - | - |
| SF - 36 Physical Functioning | 32.6±21.3 | 93.8±11.7 | <0.001 |
| CFQ total | 27.2±3.4 | 10.7±4.7 | <0.001 |

### Brain volumetry

Regional brain volumetry identified a significantly reduced brainstem volume, including the pons, in ME/CFS patients compared with healthy controls (β = −0.10% of eTIV, 95% CI −0.18 to −0.02; p = 0.013), which remained significant after Benjamini-Hochberg correction across the three tested regions (FDR-p = 0.039; **Figure 3**, **Table 2**). In contrast, deep grey matter volume and whole-brain parenchymal fraction did not differ between groups (both FDR-p = 0.98), indicating that the volumetric reduction was specific to the brainstem rather than reflecting global atrophy.

**Figure 3.**
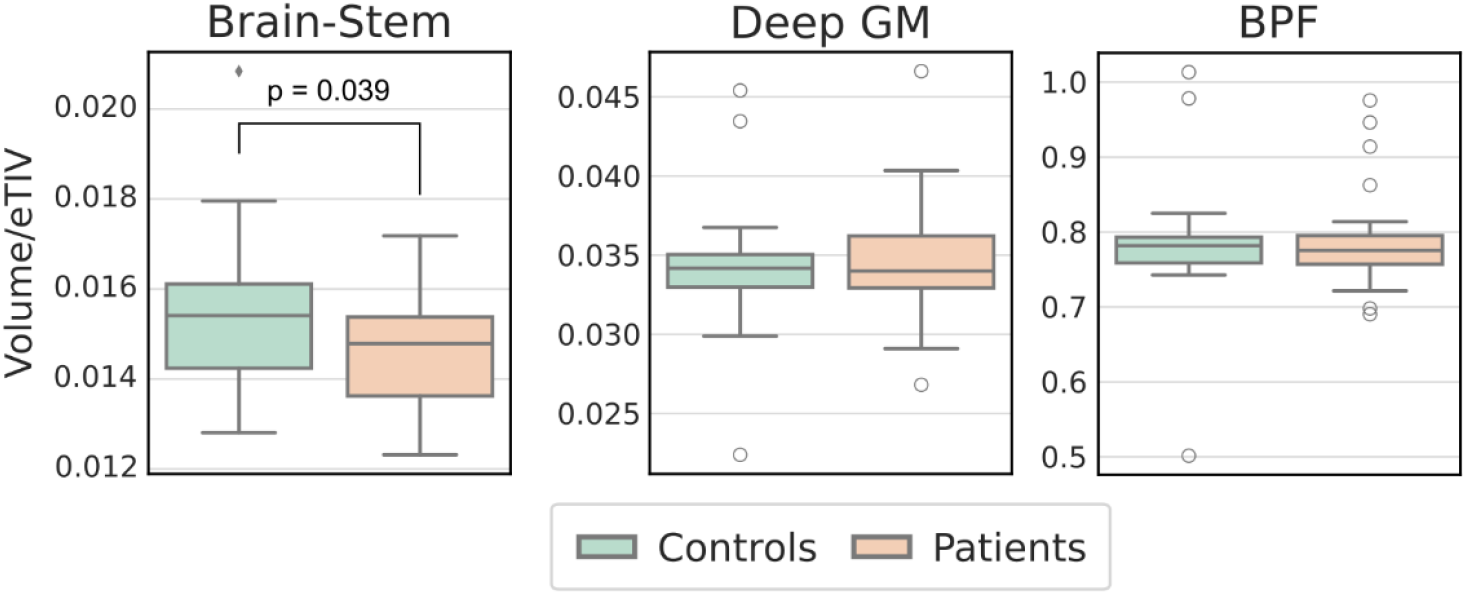
Regional volumetric measures. Boxplots of regional brain volumes in controls and patients for Brain-Stem, Deep GM and whole-brain parenchymal fraction (BPF) normalized to the estimated total intracranial volume (eTIV). <u>Abbrevations</u>: eTIV=estimated total intracranial volume, GM=grey matter, BPF=whole-brain parenchymal fraction.

**Table 2.**
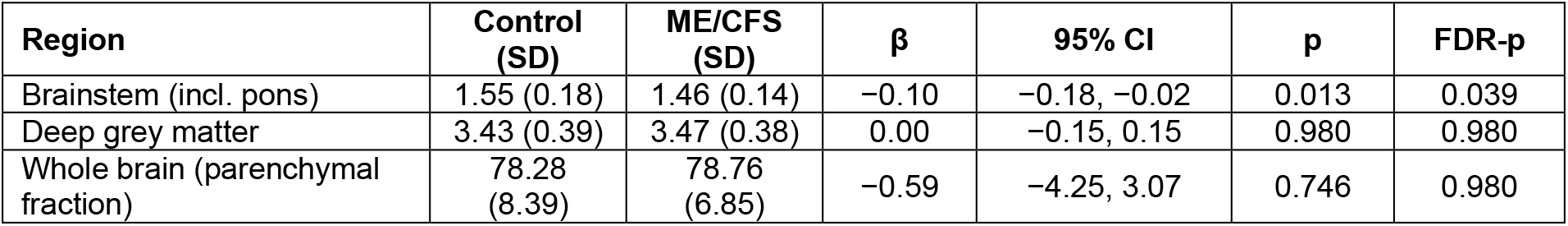
Group differences in regional brain volumes between ME/CFS patients and controls. Regional brain volumes are expressed as % of estimated total intracranial volume (eTIV); values are mean (SD). β denotes the adjusted group difference (ME/CFS minus control, in % of eTIV) estimated from linear models (volume ∼ group + age + sex + eTIV, n=53). p-values were corrected across the three tested regions using the Benjamini-Hochberg false-discovery-rate procedure (FDR-p). The brainstem measure includes the pons. *<u>Abbreviations:</u>* eTIV=estimated total intracranial volume; SD=standard deviation; CI=confidence interval; FDR=false-discovery rate.

| Region | Control (SD) | ME/CFS (SD) | $\beta$ | 95% CI | p | FDR-p |
| --- | --- | --- | --- | --- | --- | --- |
| Brainstem (incl. pons) | 1.55 (0.18) | 1.46 (0.14) | -0.10 | -0.18, -0.02 | 0.013 | 0.039 |
| Deep grey matter | 3.43 (0.39) | 3.47 (0.38) | 0.00 | -0.15, 0.15 | 0.980 | 0.980 |
| Whole brain (parenchymal fraction) | 78.28 (8.39) | 78.76 (6.85) | -0.59 | -4.25, 3.07 | 0.746 | 0.980 |

### Brain Cerebral Blood Flow

Cerebral blood flow (CBF) was quantified to characterize cerebrovascular responses to normobaric hypoxia in ME/CFS patients and healthy controls. Representative relative CBF maps acquired under normoxic and hypoxic conditions from a healthy control and a ME/CFS patient are shown in **Figure 2b**. Boxplots and spaghetti plots of regional CBF changes across normoxic and hypoxic conditions for controls and ME/CFS patients are shown in **Figure 4**. Spaghetti plots indicate greater inter-individual variability in the CBF response to hypoxia in ME/CFS patients compared with the more uniform increase observed in controls. To formally test this observation, inter-individual variability of the regional CBF response was compared between groups at each hypoxic challenge. At H_1_, patient-to-control variance ratios were elevated across all four ROIs (2.27-6.94) and the F-test was significant in 3 of 4 regions after FDR correction. These differences were absent at H_2_ (all F-test FDR p ≥ 0.848) (**Table 3**).

**Figure 4.**
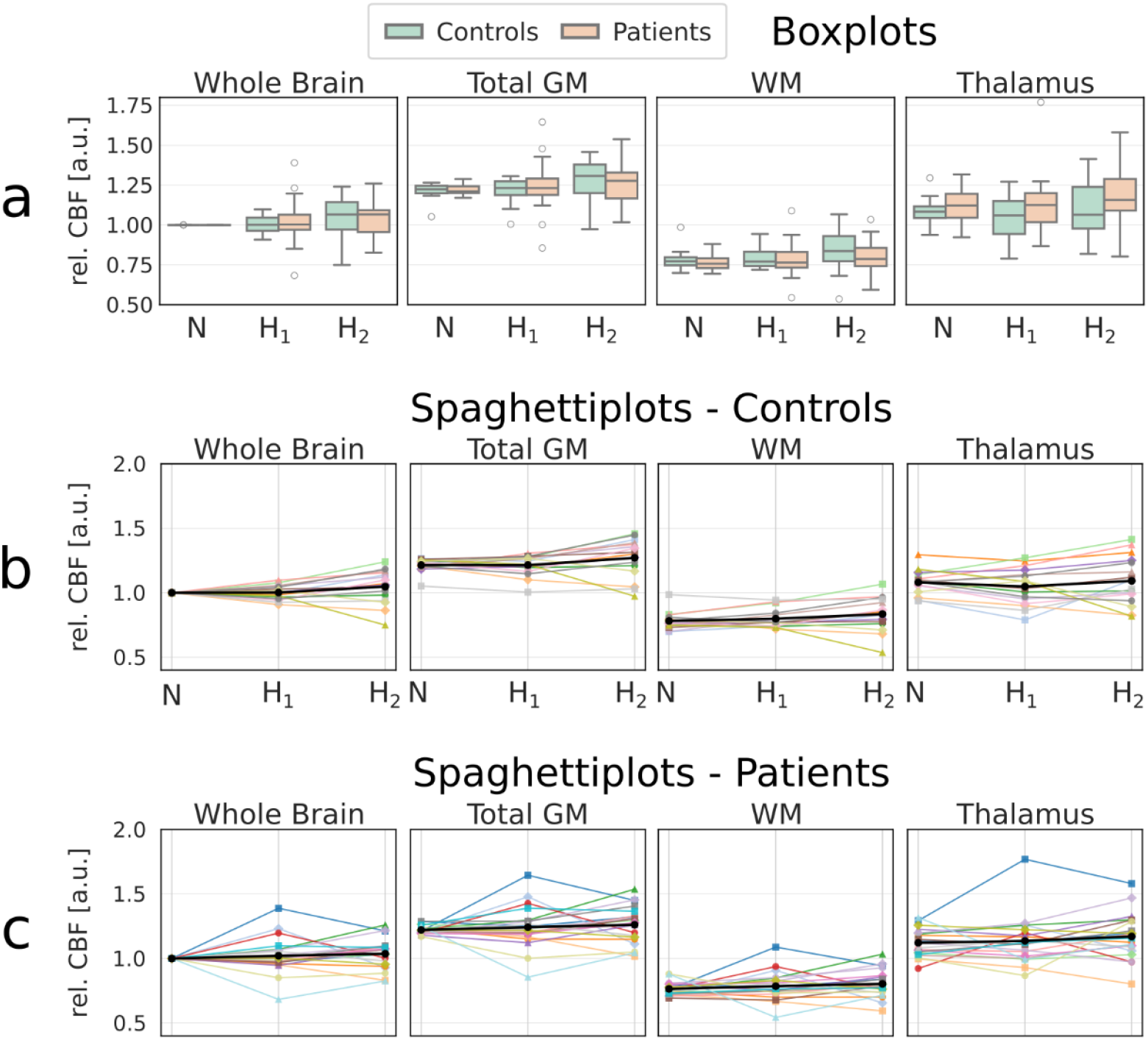
Regional cerebral blood flow response to hypoxia. Regional cerebral blood flow changes across normoxia (N) and hypoxic conditions (H1, H2) in controls and ME/CFS patients. (a) Boxplots of relative CBF values across different brain regions. (b) Spaghetti plots and group means for controls. (c) Spaghetti plots and group means for ME/CFS patients. CBF values of all conditions are normalized to the corresponding normoxic mean whole brain CBF value for each individual subject. <u>Abbreviations:</u> CBF=cerebral blood flow, GM=grey matter, WM=white matter, N=normoxia, H1=first hypoxic session, H2=second hypoxic session.

**Table 3.** Between-group dispersion of the regional CBF response to hypoxia. Between-group differences in the variability of regional cerebral blood flow responses during hypoxia. Variance estimates were calculated separately for patients and controls, and variance ratios are reported as patient-to-control variance ratios. Group differences in variance were assessed using an F-test. Both raw and false discovery rate (FDR)-corrected p-values are reported. <u>Abbreviations:</u> ROI=region of interest; GM=grey matter; H1=first hypoxic session; H2=second hypoxic session; P/C=patient-to-control variance ratio; FDR=false discovery rate.

| ROI | H1 var ratio (P/C) | H1 F p (raw) | H1 F p (FDR) | H2 var ratio (P/C) | H2 F p (raw) | H2 F p (FDR) |
| --- | --- | --- | --- | --- | --- | --- |
| Whole brain | 6.39 | <0.001 | **0.001** | 0.80 | 0.636 | 0.848 |
| White matter | 5.02 | 0.003 | **0.003** | 0.83 | 0.680 | 0.848 |
| Total GM | 6.94 | <0.001 | **0.001** | 0.92 | 0.847 | 0.848 |
| Thalamus | 2.27 | 0.110 | 0.110 | 0.74 | 0.522 | 0.848 |

H_2_ produced a consistent increase in CBF across multiple compartments in both groups. Mean whole-brain CBF increased by 4.8±13.0% in controls and 3.7±11.7% in ME/CFS patients relative to normoxia, whereas H_1_ changes were modest (0.2±5.5% and 2.2±14.0%, respectively), see **Figure 2b**. In the primary linear mixed-effects model (*CBF ∼ Condition + Group + age + sex*), significant positive effects of the second hypoxic challenge (H_2_, relative to normoxia) were observed for whole-brain CBF (β=0.042, SE=0.019, z=2.168, p=0.044), WM (β=0.045, SE=0.017, z=2.613, p=0.036), and total GM (β=0.048, SE=0.022, z=2.136, p=0.044) after Benjamini-Hochberg FDR correction across the four tested regions (whole brain, total GM, WM, and thalamus). No significant hypoxia effect was detected in the thalamus. Group (ME/CFS vs. control) effects were non-significant across all regions. Additional linear mixed-effects models including SF-36 clinical scores (physical functioning, role limitations-physical, energy/fatigue, emotional well-being; z-scored) did not attenuate hypoxia effects and contributed minimal additional variance.

### Brain Spectroscopy

Regional metabolite levels were quantified to characterize metabolic responses to normobaric hypoxia in ME/CFS patients and healthy controls. Thalamic Lac/tCr was higher in ME/CFS patients than controls at baseline (0.171 vs 0.135, ≈ 27% higher; mixed-effects group effect β=0.035, p=0.021; Welch p=0.001; **Figure 2c, 5**).

**Figure 5.**
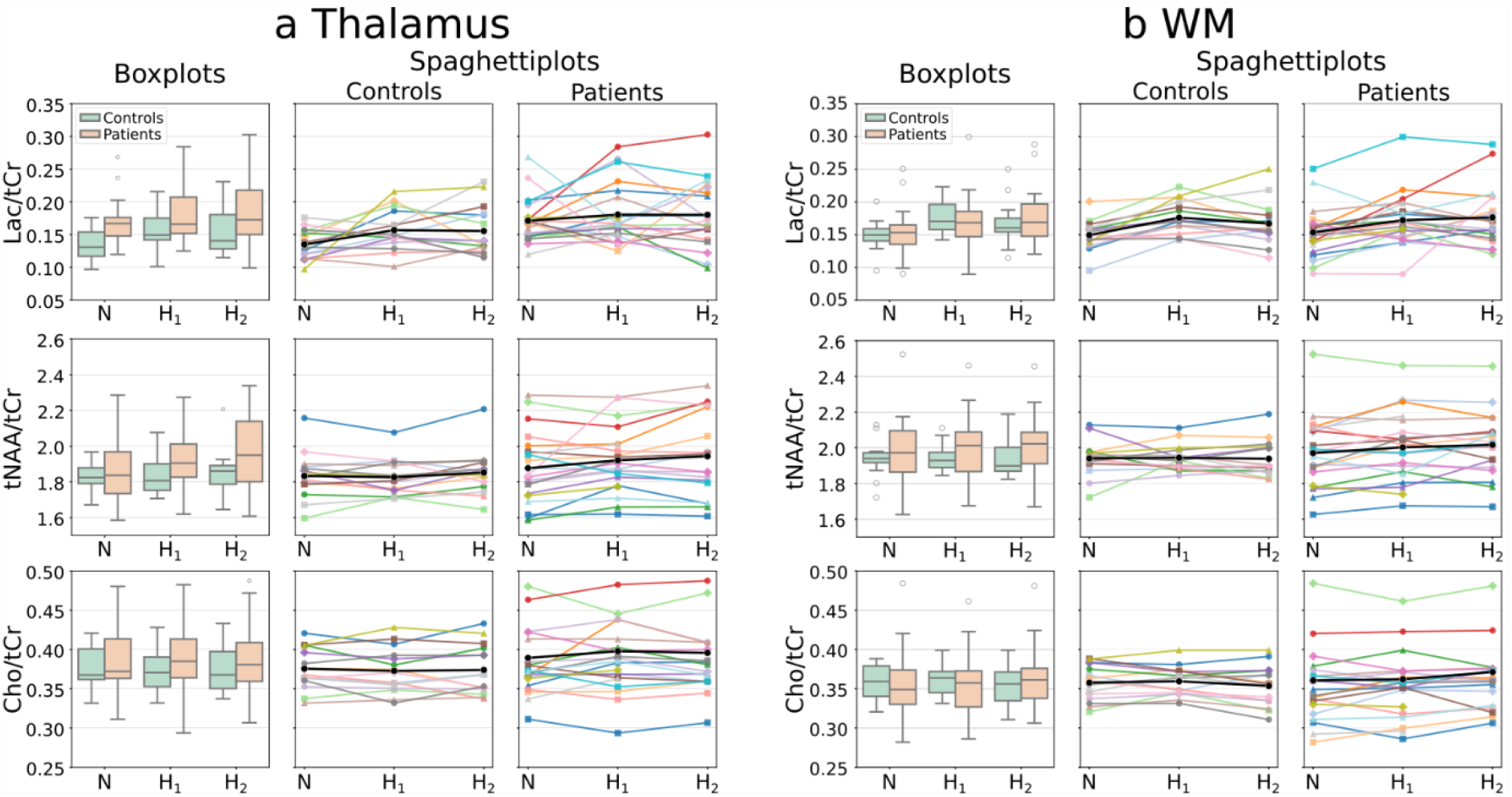
Thalamic and WM metabolite ratios during normoxia and hypoxia in ME/CFS patients and healthy controls. Metabolite ratios across normoxic (N) and hypoxic conditions (H1, H2) in the thalamus (a) and white matter (WM) (b) for controls and ME/CFS patients. Boxplots show group distributions of Lac/tCr, tNAA/tCr, and Cho/tCr ratios. Spaghetti plots illustrate individual subject trajectories together with group mean trajectories for controls and patients. Metabolite ratios were normalized to total creatine (tCr). <u>Abbreviations:</u> WM=white matter; Lac=lactate; tCr=total creatine; tNAA=total N-acetylaspartate; Cho=choline-containing compounds; N=normoxia; H1=first hypoxic session; H2=second hypoxic session.

In the primary linear mixed-effects model (*Lac/tCr ∼ Condition × Group + age + sex*), which adjusted for age and sex, this was the only metabolite to show a significant group difference after Benjamini-Hochberg FDR correction across the two ROIs, with the effect confined to the thalamus, as white-matter Lac/tCr did not differ between groups (β=-0.004, p=0.78).

Post hoc within-group analyses were performed by fitting separate linear mixed-effects models (*Lac/tCr ∼ Condition + age + sex*) for controls and ME/CFS patients. These analyses showed a hypoxia-induced increase in thalamic Lac/tCr in controls (H_1_: p=0.028; H_2_: p=0.039), but not in ME/CFS patients (H_1_: p=0.38; H_2_: p=0.46).

Neither thalamic nor white-matter tNAA/tCr differed between groups (thalamus: β=0.063, p=0.21; white matter: β=0.067, p=0.21). In both regions, tNAA/tCr declined with age (thalamus: β=-0.096, p < 0.001; white matter: β=-0.053, p=0.021). Similarly, neither thalamic nor white-matter Cho/tCr differed between groups (thalamus: β=0.020, p=0.53; white matter: β=0.010, p=0.54), and no significant hypoxia-induced changes were observed.

### Correlation network analysis

To generate hypotheses about multimodal relationships that single-variable testing may overlook, we performed an exploratory, data-driven network analysis combining regional brain volumes (eTIV-corrected), thalamic and white-matter metabolites, regional CBF, age, and clinical scores (SF-36 physical functioning and Chalder fatigue scale).

Within ME/CFS patients, Spearman correlations were estimated separately at each condition (normoxia, hypoxia 1 and 2) and their reproducibility was assessed by subject-level bootstrapping, consistent with the hypothesis-generating aim.

At normoxia a single coherent, reproducible module linked thalamic and white-matter tNAA/tCr, thalamic CBF and WM CBF, and age (bootstrap co-membership ≈ 63%, red node in **Figure 6**). Within, tNAA/tCr was highly coherent across regions (thalamic ↔ WM r=+0.89, reproducible in 100% of bootstraps) and inversely related to CBF (thalamic tNAA/tCr ↔ thalamic CBF r=-0.68, 96%; ↔ WM CBF r=-0.59, 89%), while age tracked higher white-matter CBF (r=+0.75, 98%) and lower thalamic tNAA/tCr (r=-0.66, 93%). Lactate (Lac/tCr) and the clinical scores fell outside this module (**Figure 6 and 7**).

**Figure 6.**
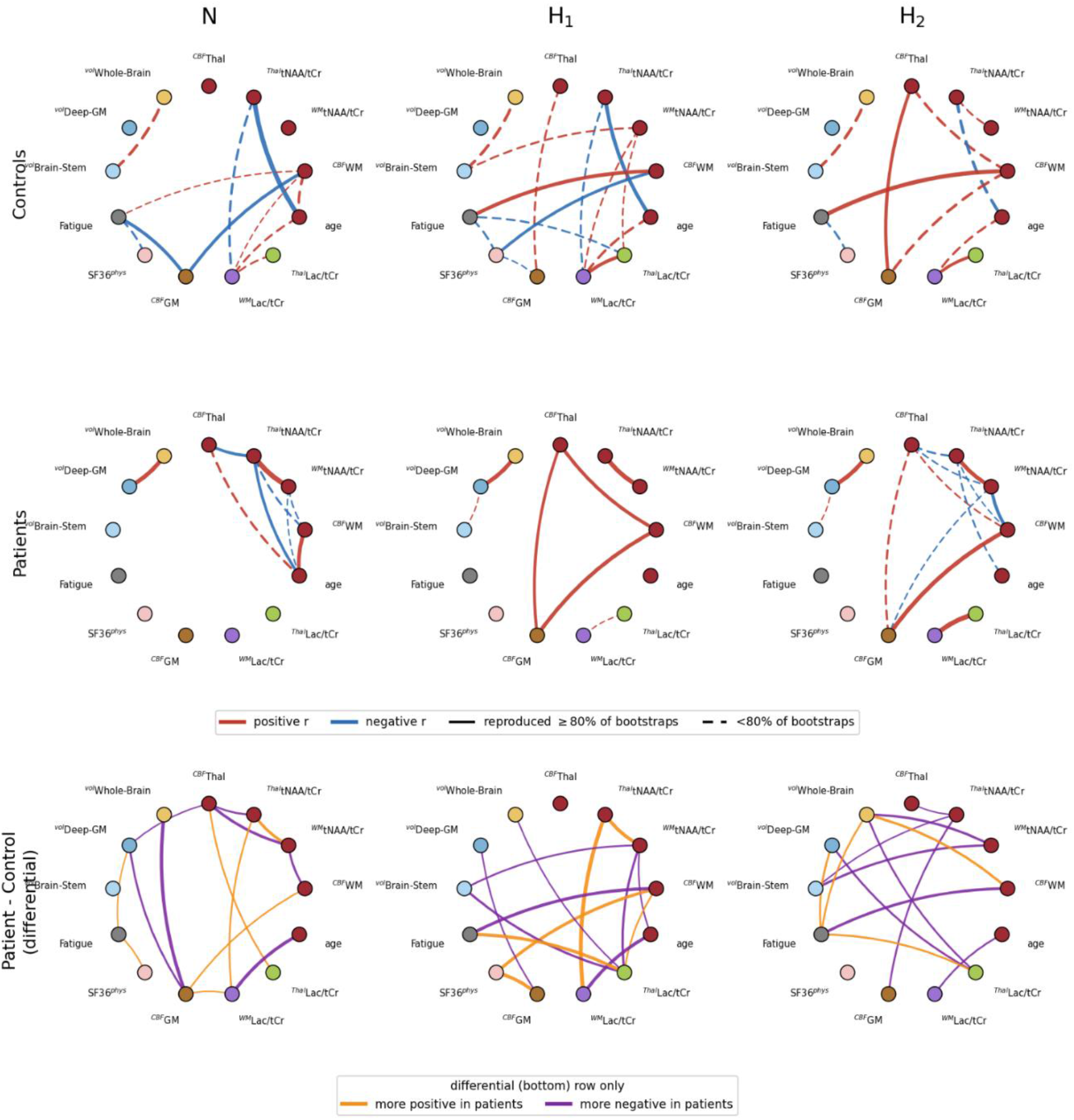
Exploratory multimodal correlation networks across conditions. Data-driven correlation networks computed within healthy controls (top row), ME/CFS patients (middle row), and as a patient-minus-control differential network (bottom row), at normoxia (N) and the first (H1) and second (H2) hypoxic challenge (columns). Nodes represent 13 variables: eTIV-corrected regional volumes (brainstem [incl. pons], deep GM, whole brain), thalamic and white-matter metabolite ratios (Lac/tCr, tNAA/tCr), regional cerebral CBF (total GM, WM, thalamus), age, and clinical scores (SF-36 physical functioning, Chalder fatigue). Node color denotes the data-driven module obtained by agglomerative clustering of the baseline patient correlation matrix; the node layout is identical in all panels to allow direct comparison. In the control and patient networks, edges are shown for |r| ≥ 0.5, with red and blue denoting positive and negative correlations, line width proportional to |r|, and solid versus dashed edges indicating correlations reproduced in ≥80% versus <80% of subject-level bootstrap resamples. In the differential networks, edges are shown for |Δr| ≥ 0.5, with orange and purple denoting relationships more positive and more negative, respectively, in patients. Patients, n=20 (N), 20 (H1) and 18 (H2); controls, n=14. Analyses are exploratory and edge reliability was assessed by bootstrap resampling. <u>Abbreviations:</u> ^vol^Brain-Stem=brainstem volume (incl. pons), ^vol^Deep-GM=deep grey-matter volume, ^vol^Whole-Brain=whole-brain volume (parenchymal fraction), Δr=between-group difference in correlation (patient - control), eTIV=estimated total intracranial volume, Fatigue=Chalder fatigue total score, GM=grey matter, H1=first hypoxic challenge, H2=second hypoxic challenge, Lac/tCr=lactate-to-total-creatine ratio, ^Thal^Lac/tCr=thalamic Lac/tCr, ^WM^ Lac/tCr=white-matter Lac/tCr, ME/CFS=myalgic encephalomyelitis/chronic fatigue syndrome, N=normoxia, ^Thal^tNAA/tCr=thalamic tNAA/tCr, ^WM^tNAA/tCr=white-matter tNAA/tCr, ^CBF^GM=total grey-matter CBF, ^CBF^Thal=thalamic CBF, ^CBF^WM=white-matter CBF, r=Spearman correlation coefficient, SF-36=36-item Short Form Health Survey, ^physical^SF36=SF-36 physical-functioning subscale, tNAA/tCr=total N-acetylaspartate-to-total-creatine ratio, WM=white matter.

**Figure 7.**
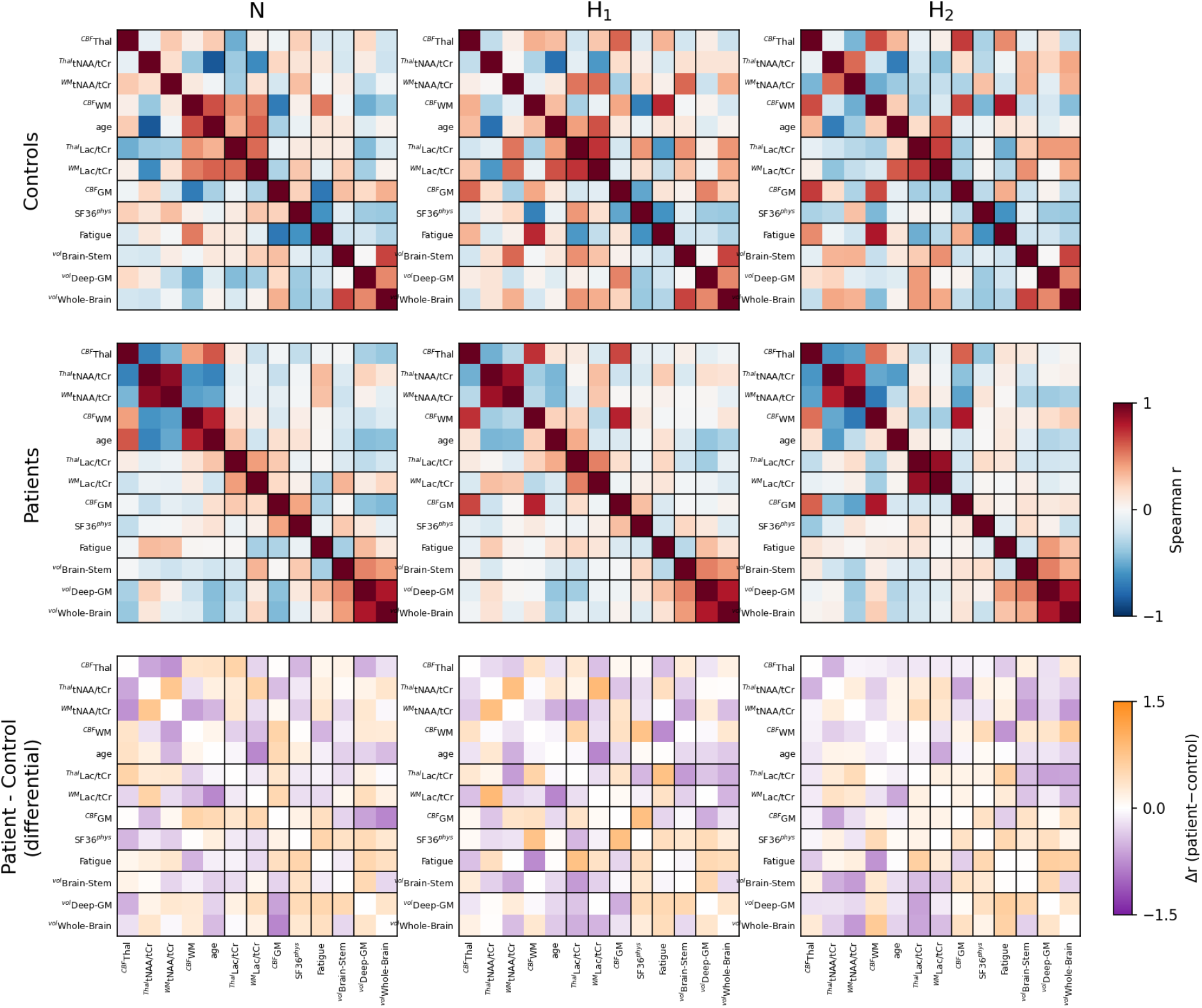
Clustered correlation matrices across conditions. Spearman correlation matrices among the same 13 variables (Figure 5) within healthy controls (top row), ME/CFS patients (middle row), and as the patient-minus-control difference (Δr; bottom row), at normoxia (N) and the first (H1) and second (H2) hypoxic challenge (columns). In every panel, variables are arranged in a fixed order defined by the data-driven modules of the baseline patient matrix, and black lines mark the module boundaries. For the control and patient rows, colour encodes Spearman r (scale −1 to +1); for the differential row, colour encodes Δr=r(patient) - r(control) (scale −1.5 to +1.5); red and blue denote positive and negative Spearman r in the control and patient rows, and orange and purple denote relationships more positive and more negative in patients in the differential row. Patients, n=20 (N), 20 (H1) and 18 (H2); controls, n=14. Analyses are exploratory. <u>Abbreviations</u>: ^vol^Brain-Stem=brainstem volume (incl. pons), ^vol^Deep-GM=deep grey-matter volume, ^vol^Whole-Brain=whole-brain volume (parenchymal fraction), Δr=between-group difference in correlation (patient - control), eTIV=estimated total intracranial volume, Fatigue=Chalder fatigue total score, GM=grey matter, H1=first hypoxic challenge, H2=second hypoxic challenge, Lac/tCr=lactate-to-total-creatine ratio, ^Thal^Lac/tCr=thalamic Lac/tCr, ^WM^ Lac/tCr=white-matter Lac/tCr, ME/CFS=myalgic encephalomyelitis/chronic fatigue syndrome, N=normoxia, ^Thal^tNAA/tCr=thalamic tNAA/tCr, ^WM^tNAA/tCr=white-matter tNAA/tCr, ^CBF^GM=total grey-matter CBF, ^CBF^Thal=thalamic CBF, ^CBF^WM=white-matter CBF, r=Spearman correlation coefficient, SF-36=36-item Short Form Health Survey, ^physical^SF36=SF-36 physical-functioning subscale, tNAA/tCr=total N-acetylaspartate-to-total-creatine ratio, WM=white matter.

In the differential (patient - control) network, three patterns stood out: the whole-brain volume ↔ GM CBF relationship reversed in sign (^vol^Whole-Brain↔^CBF^GM: control r=+0.36, patient r=-0.44); the thalamic tNAA/tCr-thalamic CBF coupling was strongly negative in patients but weakly negative in controls (r=-0.68 vs −0.12); and thalamic and white-matter tNAA/tCr were far more tightly correlated in patients (r=+0.89 vs +0.17).

Across conditions, the tNAA/tCr-CBF-age module was preserved under both hypoxic challenges, whereas in patients the thalamic Lac/tCr↔WM Lac/tCr coupling strengthened under hypoxia, peaking at the second challenge (r=+0.42, +0.50 and +0.86 across normoxia, H_1_ and H_2_; in controls it was already strong at baseline, r=+0.57) (**Figure 6 and 7**).

## Discussion

This proof-of-concept multimodal neuroimaging study performed structural volumetry, arterial-spin-labelling imaging, and proton magnetic resonance spectroscopy under controlled normoxic and two hypoxic conditions in ME/CFS patients and matched controls. We found preserved gross CBF reactivity to hypoxia yet, in patients, greater initial inter-individual variability in the CBF response and no significant group difference in its mean. We also found elevated baseline thalamic Lac/tCr in patients, that failed to respond to hypoxia; selective brainstem reduction; and, in exploratory network analyses, a regionally specific inverse coupling between thalamic tNAA/tCr and CBF in patients. While these findings are preliminary and the hypoxic challenge not equivalent to physical exertion, the experimental findings of our study support the concept of impaired neuro-metabolic and vascular adaptive capacity in ME/CFS.

Previous neuroimaging studies in ME/CFS have largely examined single modalities and have been heterogeneous, reporting modest grey- or white-matter changes, subtle hypoCBF, variable metabolite findings, or no abnormality^17–20^. Our results nonetheless converge with, and integrate, several specific strands of this literature. The elevated baseline thalamic Lac/tCr is consistent with the most reproducible spectroscopic finding in ME/CFS - increased ventricular and cerebral lactate on ¹H-MRS, which has been linked to reduced cortical glutathione, and thus to oxidative stress and a glycolytic shift, supporting a bioenergetic interpretation^6,7,21,22^. The inverse thalamic tNAA/tCr to white-matter CBF coupling (r=-0.59 in patients) and the marginal white-matter hypoCBF are in keeping with reports of reduced CBF^11^ and of neuroinflammation and mitochondrial dysfunction^8,10^. Nevertheless, it should be noted that reduced ASL-derived white-matter CBF may also indicate later label arrival, reflecting reduced vascular efficiency.

The selective brainstem volume reduction adds to repeated evidence of brainstem structural and functional abnormality and its links to autonomic dysregulation and fatigue^23,24^, although the reported direction of brainstem volume change has been inconsistent across cohorts.

Beyond these single-modality observations, our combined acquisition links a regionally specific resting metabolic abnormality to its vascular and structural context within the same participants. In exploratory analyses, the altered thalamic tNAA/tCr-CBF coupling in patients is, tentatively, consistent with the impaired cerebral metabolic flexibility proposed in the post-COVID and ME/CFS overlap literature^25,26^ and warrants confirmation in larger cohorts.

The elevation of thalamic lactate despite normal arterial oxygenation is consistent with the “virtual hypoxia” concept, in which increased metabolic demand or impaired substrate utilisation produces a functionally hypoxic tissue state. Together with the inverse thalamic tNAA/tCr-CBF coupling, this resting pattern was the principal between-group difference; we did not measure markers of inflammation or oxidative stress and thus cannot establish the upstream mechanism. However, this pattern is consistent with intrinsic mitochondrial injury or bioenergetic inefficiency, whereby the brain behaves as if it is already operating under chronic oxygen limitation^27^.

If validated in independent cohorts, the resting thalamic Lac/tCr elevation and the thalamic tNAA/tCr-CBF coupling could be explored as candidate stratification markers for trials of interventions targeting tissue oxygenation or mitochondrial function. Recent work^28^ demonstrated symptomatic benefit from Hyperbaric Oxygen Therapy (HBOT) in a subset of ME/CFS and long-COVID patients, although response heterogeneity remains high. Such a biomarker-guided approach is hypothesis-generating and would require prospective validation.

Limitations of this study include the modest sample size and cross-sectional design. Because the hypoxic challenge probes neurovascular and metabolic reactivity, but is not equivalent to physical exertion, our findings cannot be directly attributed to post-exertional malaise. Our correlation-based coupling analyses were exploratory: they were performed within relatively small group- and condition-specific subsamples, used subject-level bootstrap reproducibility in place of the family-wise multiple-comparison correction applied to the primary volumetric, perfusion and spectroscopic analyses (Benjamini-Hochberg FDR). Across modalities we sought to keep the analyzed regions as consistent as possible and deliberately favored large, well-defined compartments, so as to limit spurious associations in this modest sample; the volumetric nodes were accordingly restricted to those of the primary analysis (brainstem, deep grey matter and whole brain), with the thalamus subsumed within deep grey matter rather than modelled as a separate node. This decision also reflects the comparatively limited reliability of arterial-spin-labelling (ASL) CBF in small, deep structures such as the thalamus and brainstem, in which long and variable arterial transit times and low signal-to-noise ratio reduce measurement stability^29^, so that edges involving these regions should be interpreted with corresponding caution. WM and thalamic spectroscopic voxels were, in addition, positioned and analyzed to minimize cerebrospinal-fluid partial-volume contamination, further guarding against artefactual coupling.

Here presented results should be regarded as hypothesis-generating, and the apparent group difference in hypoxia-induced lactate-volume coupling in particular warrants confirmation in larger, adequately powered cohorts. In addition, spectroscopic measurements were limited to a 2D acquisition with relatively large voxel sizes, resulting in substantial point spread function effects and partial volume contamination. Consequently, contributions from surrounding cerebrospinal fluid (CSF) to the measured lactate signal cannot be fully excluded. Future work will focus on implementing more advanced spectroscopic acquisitions, including a 3D semi-LASER sequence with concentric ring trajectory sampling, enabling substantially smaller voxel size, improved spatial coverage, and reduced partial volume effects^30^. In addition, it should be mentioned that interpretation of thalamic perfusion should be made with caution, as the mixed grey- and white-matter composition of the thalamus increases susceptibility to segmentation inaccuracies and partial volume effects.

Nevertheless, the convergence of CBF dynamics, spectroscopic shifts, volumetric selectivity, and altered correlation plasticity provides a coherent mechanistic framework. Finally, although the SF-36 was used in line with most studies in the field, it has not been formally validated in ME/CFS and does not capture all aspects of the disease; condition-specific instruments may better characterize the clinical phenotype in future work. Larger, longitudinal studies incorporating symptom provocation and targeted interventions will be essential to translate these proof-of-concept insights into clinically actionable biomarkers.

This proof-of-concept multimodal study identified a regionally specific resting metabolic abnormality in ME/CFS, namely elevated baseline thalamic Lac/tCr, together with a selective brainstem volume reduction, an altered thalamic tNAA/tCr-CBF relationship and a hypoxia-dependent strengthening of thalamic-white-matter lactate coupling. These preliminary findings inform us on a potential bioenergetic / “virtual hypoxia” component in the pathogenesis of ME/CFS and warrant validation as metrics for patient stratification and treatment monitoring.

## Data Availability

All data produced in the present study are available upon reasonable request from the corresponding authors

## Acknowledgements

This work was supported by the Vienna Science and Technology Grant (ME-CFS24-018 to LH, TZ and KK) and the We&Me Foundation (MRI in ME/CFS to LH). Additionally, this work was supported by the Austrian Science Fund (10.55776/KLI1106, 10.55776/KLP1326625) to FN. We extend our warmest thanks to the WE & Me Foundation and the supportive framework it provides for the generous guidance, thoughtful feedback, and steady encouragement that shaped this study. We are especially grateful to the individuals with lived experience of ME/CFS who kindly shared their insights and perspectives, and to the patients and controls who generously took the time and effort to participate in the study.

## Author contributions

**Conceptualization:** L.H. and K.K. **Methodology:** V.B., L.H. and K.K. **Software:** V.B. **Formal analysis:** V.B. and L.H. **Investigation:** K.E., K.K. and L.H. **Data curation:** V.B., K.E., K.K. and L.H. **Visualization:** V.B. and L.H. **Writing – original draft:** V.B., L.H. and K.K. **Writing – review & editing:** All authors. **Supervision:** F.N. and L.H. **Funding acquisition:** L.H., K.K. and F.N. All authors reviewed and approved the final version of the manuscript.

## COI

**Viola Bader**: No conflicts of interest

**Katharina Estermann**: No conflicts of interest

**Eva Niess**: No conflicts of interest

**Tobias Zrzavy**: participated in meetings sponsored by or received travel funding from Biogen, Merck, Novartis, Roche, Sanofi and Teva; None are relevant to this publication

**Florian Fischmeister**: No conflicts of interest

**Teresa Haider**: No conflicts of interest

**Birgit Ludwig**: reports a relationship with Medical Chamber of Vienna that includes: funding grants; None are relevant to this publication.

**Frederik Barkhof**: reports serving on steering committees and Data Safety Monitoring Boards for Biogen, Merck, ATRI/ACTC, and Prothena; consulting fees from Roche, Celltrion, Rewind Therapeutics, Merck, IXICO, Janssen, and Combinostics; research agreements with Merck, Biogen, GE Healthcare, and Roche; and is a co-founder and shareholder of Queen Square Analytics Ltd.; None are relevant to this publication.

**Henk JMM Mutsaerts**: received payments for consulting from TheriniBio; None are relevant to this publication.

**Gregor Kasprian**: Speaker’s Bureau: Alexion; Philips. Other Financial or Material Support:

BRACCO, Lilly; None are relevant to this publication.

**Fabian Niess**: No conflicts of interest

**Wolfgang Bogner**: No conflicts of interest

**Kathrin Kollndorfer**: No conflicts of interest

**Lukas Haider**: received payments for consulting, speaking or travel, from Eisai, Lilly, Roche, Siemens and Teva; None are relevant to this publication.

## References

1. Institute of Medicine (US) Committee on the Diagnostic Criteria for Myalgic Encephalomyelitis/Chronic Fatigue Syndrome. Beyond Myalgic Encephalomyelitis/Chronic Fatigue Syndrome: Redefining an Illness 2015. National Academies Press; 2015. Accessed May 4, 2026. https://www.nationalacademies.org/publications/19012

2. Mahad DJ, Ziabreva I, Campbell G, et al. Mitochondrial changes within axons in multiple sclerosis. Brain. 2009;132(5):1161–1174. doi:10.1093/brain/awp046

3. Mahad DH, Trapp BD, Lassmann H. Pathological mechanisms in progressive multiple sclerosis. Lancet Neurol. 2015;14(2):183–193. doi:10.1016/S1474-4422(14)70256-X

4. Attwell D, Laughlin SB. An Energy Budget for Signaling in the Grey Matter of the Brain. J Cereb Blood Flow Metab. 2001;21(10):1133–1145. doi:10.1097/00004647-200110000-00001

5. Trapp BD, Stys PK. Virtual hypoxia and chronic necrosis of demyelinated axons in multiple sclerosis. Lancet Neurol. 2009;8(3):280–291. doi:10.1016/S1474-4422(09)70043-2

6. Tomas C, Brown A, Strassheim V, Elson J, Newton J, Manning P. Cellular bioenergetics is impaired in patients with chronic fatigue syndrome. PLOS ONE. 2017;12(10):e0186802. doi:10.1371/journal.pone.0186802

7. Naviaux RK, Naviaux JC, Li K, et al. Metabolic features of chronic fatigue syndrome. Proc Natl Acad Sci U S A. 2016;113(37):E5472–5480. doi:10.1073/pnas.1607571113

8. Nakatomi Y, Mizuno K, Ishii A, et al. Neuroinflammation in Patients with Chronic Fatigue Syndrome/Myalgic Encephalomyelitis: An 11C-(R)-PK11195 PET Study. Journal of Nuclear Medicine. 2014;55(6):945–950. doi:10.2967/jnumed.113.131045

9. Montoya JG, Holmes TH, Anderson JN, et al. Cytokine signature associated with disease severity in chronic fatigue syndrome patients. Proceedings of the National Academy of Sciences. 2017;114(34):E7150–E7158. doi:10.1073/pnas.1710519114

10. Lee JS, Sato W, Son CG. Brain-regional characteristics and neuroinflammation in ME/CFS patients from neuroimaging: A systematic review and meta-analysis. Autoimmunity Reviews. 2024;23(2):103484. doi:10.1016/j.autrev.2023.103484

11. van Campen CLMC, Verheugt FWA, Rowe PC, Visser FC. Cerebral blood flow is reduced in ME/CFS during head-up tilt testing even in the absence of hypotension or tachycardia: A quantitative, controlled study using Doppler echography. Clin Neurophysiol Pract. 2020;5:50–58. doi:10.1016/j.cnp.2020.01.003

12. Alsop DC, Detre JA, Golay X, et al. Recommended implementation of arterial spin-labeled perfusion MRI for clinical applications: A consensus of the ISMRM perfusion study group and the European consortium for ASL in dementia. Magnetic Resonance in Medicine. 2015;73(1):102–116. doi:10.1002/mrm.25197

13. Wijnen JP, van Asten JJA, Klomp DWJ, et al. Short echo time 1H MRSI of the human brain at 3T with adiabatic slice-selective refocusing pulses; reproducibility and variance in a dual center setting. J Magn Reson Imaging. 2010;31(1):61–70. doi:10.1002/jmri.21999

14. Kreis R. The trouble with quality filtering based on relative Cramér-Rao lower bounds. Magn Reson Med. 2016;75(1):15–18. doi:10.1002/mrm.25568

15. Barnden LR, Shan ZY, Staines DR, et al. Intra brainstem connectivity is impaired in chronic fatigue syndrome. Neuroimage Clin. 2019;24:102045. doi:10.1016/j.nicl.2019.102045

16. Barnden LR, Crouch B, Kwiatek R, et al. A brain MRI study of chronic fatigue syndrome: evidence of brainstem dysfunction and altered homeostasis. NMR in Biomedicine. 2011;24(10):1302–1312. doi:10.1002/nbm.1692

17. Li X, Julin P, Li TQ. Limbic Perfusion Is Reduced in Patients with Myalgic Encephalomyelitis/Chronic Fatigue Syndrome (ME/CFS). Tomography. 2021;7(4):675–687. doi:10.3390/tomography7040056

18. Walitt B, Singh K, LaMunion SR, et al. Deep phenotyping of post-infectious myalgic encephalomyelitis/chronic fatigue syndrome. Nat Commun. 2024;15(1):907. doi:10.1038/s41467-024-45107-3

19. Godlewska BR, Sylvester AL, Emir UE, et al. Brain and muscle chemistry in myalgic encephalitis/chronic fatigue syndrome (ME/CFS) and long COVID: a 7T magnetic resonance spectroscopy study. Mol Psychiatry. 2025;30(11):5215–5226. doi:10.1038/s41380-025-03108-8

20. Shan ZY, Barnden LR, Kwiatek RA, Bhuta S, Hermens DF, Lagopoulos J. Neuroimaging characteristics of myalgic encephalomyelitis/chronic fatigue syndrome (ME/CFS): a systematic review. J Transl Med. 2020;18(1):335. doi:10.1186/s12967-020-02506-6

21. Shankar V, Wilhelmy J, Curtis EJ, et al. Oxidative stress is a shared characteristic of ME/CFS and Long COVID. Proc Natl Acad Sci USA. 2025;122(28):e2426564122. doi:10.1073/pnas.2426564122

22. Che X, Ranjan A, Guo C, et al. Heightened innate immunity may trigger chronic inflammation, fatigue and post-exertional malaise in ME/CFS. npj Metab Health Dis. 2025;3(1):34. doi:10.1038/s44324-025-00079-w

23. Chu L, Valencia IJ, Garvert DW, Montoya JG. Onset Patterns and Course of Myalgic Encephalomyelitis/Chronic Fatigue Syndrome. Front Pediatr. 2019;7:12. doi:10.3389/fped.2019.00012

24. Yu Q, Kwiatek RA, Del Fante P, et al. Distinct white matter alteration patterns in post-infectious and gradual onset chronic fatigue syndrome revealed by diffusion MRI. Sci Rep. 2025;15(1):24256. doi:10.1038/s41598-025-09379-z

25. Schwichtenberg K, Hartung T, Heine J, et al. Association of structural brain changes with cognitive deficits and fatigue in patients with post-COVID-19 condition. Brain Communications. 2026;8(2):fcag099. doi:10.1093/braincomms/fcag099

26. Bansal AS, Seton KA, Brooks JCW, Carding SR. Cognitive Dysfunction in Myalgic Encephalomyelitis/Chronic Fatigue Syndrome—Aetiology and Potential Treatments. IJMS. 2025;26(5):1896. doi:10.3390/ijms26051896

27. Franklin RJM, ffrench-Constant C, Edgar JM, Smith KJ. Neuroprotection and repair in multiple sclerosis. Nat Rev Neurol. 2012;8(11):624–634. doi:10.1038/nrneurol.2012.200

28. Kim L, Cammà G, Peters CK, et al. Hyperbaric oxygen therapy improves clinical symptoms and functional capacity and restores thalamic connectivity in ME/CFS. Neurology. Preprint posted online October 31, 2025. doi:10.1101/2025.10.29.25339096

29. Haller S, Zaharchuk G, Thomas DL, Lovblad KO, Barkhof F, Golay X. Arterial Spin Labeling Perfusion of the Brain: Emerging Clinical Applications. Radiology. 2016;281(2):337–356. doi:10.1148/radiol.2016150789

30. (ISMRM 2025) Measuring Lactate Levels in Gray and White Matter of the healthy Human Brain using semi-LASER MRSI with Concentric Ring Trajectory Encoding at 3T. Accessed May 7, 2026. https://archive.ismrm.org/2025/0601.html

31. BASIL: A toolbox for perfusion quantification using arterial spin labelling | Imaging Neuroscience | MIT Press. Accessed May 6, 2026. https://direct.mit.edu/imag/article/doi/10.1162/imag_a_00041/118215/BASIL-A-toolbox-for-perfusion-quantification-using

32. Bullinger M, Kirchberger I, Ware J. Der deutsche SF-36 Health Survey Übersetzung und psychometrische Testung eines krankheitsübergreifenden Instruments zur Erfassung der gesundheitsbezogenen Lebensqualität. J Public Health. 1995;3(1):21–36. doi:10.1007/BF02959944

33. Bullinger M, Kirchberger I. SF-36. Fragebogen Zum Gesundheitszustand. Handanweisung. Hogrefe; 1998.

34. Chalder T, Berelowitz G, Pawlikowska T, et al. Development of a fatigue scale. Journal of Psychosomatic Research. 1993;37(2):147–153. doi:10.1016/0022-3999(93)90081-P

35. Martin A, Staufenbiel T, Gaab J, Rief W, Brähler E. Messung chronischer Erschöpfung – Teststatistische Prüfung der Fatigue Skala (FS). Zeitschrift für Klinische Psychologie und Psychotherapie. 2010;39(1):33–44. doi:10.1026/1616-3443/a000010

36. Posit team. *RStudio: Integrated Development Environment for R*. Posit Software, PBC; 2025. http://www.posit.co/

